# Regional Disparities and Temporal Trends in Rheumatic Heart Disease Burden in Nepal: A Systematic Review and Meta-Analysis

**DOI:** 10.1101/2025.09.06.25335242

**Authors:** Sandip Pandey, Apil Tiwari, Alisha Bhattarai, Anu Timalsina, Asmit Pandey, Aashis Poudel, Aakash Neupane, Deepak Jung Subedi, Kumar Bahadur Bista, Shrishty Deubhadel

## Abstract

**Introduction:** Rheumatic heart disease (RHD) is a leading cause of preventable cardiovascular disease in low- and middle-income countries. Nepal is an endemic region, but no nationally representative synthesis of prevalence data exists. We conducted a systematic review and meta-analysis to estimate RHD prevalence, identify geographic disparities, and assess certainty of evidence.

**Methods:** We searched PubMed, Embase, and Scopus for population-based studies reporting echocardiographically confirmed RHD prevalence in Nepal (January 1990 to July 2025). Pooled prevalence was estimated using a generalized linear mixed model with logit transformation. Subgroup analysis compared the Kathmandu Valley with non-Kathmandu regions. Meta-regression examined publication year, sample size, and geographic region. Certainty of evidence was assessed using GRADE. The protocol was registered with PROSPERO (CRD42025126445).

**Results:** Ten studies (209,815 participants; 5,440 cases) were included. Pooled RHD prevalence was 3.1% (95% CI 1.6% to 5.8%), with high heterogeneity (I^2^ = 99.8%). Excluding the nationwide survey, prevalence in the Kathmandu Valley was 1.1% (95% CI 0.9% to 1.3%), compared with 7.2% (95% CI 3.7% to 13.3%) outside Kathmandu (OR 6.52, 95% CI 4.83 to 8.81; p < 0.001). In univariable meta-regression, more recent studies showed higher prevalence (β = 0.072; p = 0.012) and larger studies showed lower prevalence (β = −0.87; p < 0.001). After multivariable adjustment, the temporal association became non-significant, suggesting the apparent rise reflects diagnostic upgrading and small-study effects rather than a true increase. The model explained 93.5% of between-study variance. GRADE certainty was low for overall and non-Kathmandu estimates and moderate for Kathmandu.

**Conclusion:** RHD prevalence outside Kathmandu is approximately six times higher than in urban areas. No true decline was detected over three decades after accounting for diagnostic and study-size differences. These findings support integrating RHD prevention, community-based echocardiographic screening, and equitable access to benzathine penicillin prophylaxis into Nepal’s decentralized health system.

## Introduction

Rheumatic heart disease is the chronic sequela of acute rheumatic fever caused by group A Streptococcus pharyngeal infection. An estimated 40.5 million people live with RHD globally, with 306,000 deaths attributed to the condition annually(1). The disease disproportionately affects children and young adults in low- and middle-income countries, causing irreversible valvular damage, heart failure, stroke, and premature death (2)(3). Despite advances in antibiotic availability and primary healthcare, RHD persists in settings where poverty, overcrowding, and weak health systems sustain streptococcal transmission and limit access to secondary prophylaxis.

Nepal, a country situated in the Himalayas, is recognized as an endemic region for RHD(4). Socioeconomic conditions characterised by poverty, seasonal migration, household overcrowding, and limited healthcare access create an environment conducive to group A Streptococcus transmission and progression to acute rheumatic fever and RHD(5). Published studies have reported widely varying prevalence, ranging from 1.35 to 13.2 per 100 screened children, but these estimates derive from geographically restricted, methodologically heterogeneous studies(6) (7). No synthesis has characterised the national burden, its geographic distribution across Nepal’s ecologically and socioeconomically diverse provinces, or its trajectory over time.

This evidence gap carries direct policy consequences. Nepal’s 2015 constitutional reform devolved healthcare delivery to 753 local governments and seven provinces. Without a comprehensive national picture, provincial health plans, benzathine penicillin supply chains, and echocardiographic screening programmes cannot be rationally prioritised or resourced. A pooled analysis of South Asian data suggests a 24.8% regional decline in RHD prevalence from 1991 to 2021, but whether Nepal follows or deviates from this trajectory remains unanswered(8).

We conducted a systematic review and meta-analysis to address four specific objectives: (1) to estimate the national pooled prevalence of RHD in Nepal; (2) to quantify the geographic disparity between the Kathmandu Valley and outside-Kathmandu populations; (3) to examine temporal trends in prevalence; and (4) to identify sources of heterogeneity through meta-regression, including the contribution of diagnostic method, study size, and geographic region.

## Methods

### Protocol and registration

This review was conducted in accordance with PRISMA 2020 guidelines.[9] The protocol was prospectively registered with PROSPERO (CRD42025126445). During the review, the risk-of-bias tool was amended from the Newcastle-Ottawa Scale to the Joanna Briggs Institute (JBI) checklist for prevalence studies, to better align with the included study designs. This amendment was recorded in the PROSPERO record on 6 September 2025.

### Search Strategy and study selection

This systematic review was conducted in accordance with the Preferred Reporting Items for Systematic Reviews and Meta-Analyses (PRISMA) guidelines(9). We searched PubMed, Embase, and Scopus from January 1990 to 1 July 2025, without language restriction. We also searched Nepalese institutional repositories and grey literature sources. The search strategy combined controlled vocabulary and free-text terms:

(“rheumatic heart disease” OR “RHD” OR “chronic rheumatic heart disease” OR “recurrent rheumatic fever”) AND (“Nepal” OR “Himalaya*” OR “Nepalese”)

Search results were exported to Rayyan for deduplication. Two reviewers screened titles and abstracts, then full texts, independently. Disagreements were resolved by consensus or a third reviewer.

### Eligibility Criteria

Studies were eligible if they were population-based surveys conducted in Nepal, used echocardiography to confirm RHD diagnosis (adhering to World Heart Federation [WHF] criteria or equivalent established guidelines for studies conducted before the 2012 WHF publication) (10), reported extractable prevalence data with numerator and denominator, and included at least 100 participants. We excluded case reports, review articles, interventional studies without baseline prevalence data, and studies using a patient or valvular heart disease cohort as the denominator.

### Data Extraction and Quality Assessment

Two reviewers independently extracted: first author, year, region, sample size, number of cases, prevalence, diagnostic method, and adherence to WHF criteria. Geographic classification used an operational definition: studies conducted within the Kathmandu Valley (Kathmandu, Lalitpur, and Bhaktapur districts) were classified as Kathmandu; all other province-based or community surveys were classified as non-Kathmandu, representing predominantly rural or mixed settings. This definition aligns with Nepal’s federal demographic distribution, in which the Kathmandu Valley constitutes the primary urban hub. The nationwide survey (Regmi 2023) was classified separately and excluded from geographic subgroup analysis to avoid misclassification.

Risk of bias was assessed with the JBI checklist for prevalence studies(11).

### Statistical analysis

Prevalence estimates were converted to proportions. Pooled prevalence was estimated using a generalised linear mixed model (GLMM) with logit transformation (PLOGIT), with all results back-transformed to percentages. The GLMM was chosen for its stability in low-prevalence settings and its avoidance of continuity corrections. Only random-effects models were used.

Between-study heterogeneity was quantified using Cochran’s Q test, tau-squared (tau^2^), and I^2^. Prediction intervals were calculated to reflect the expected range of prevalence in a new Nepalese setting.

Pre-specified subgroup analysis compared Kathmandu and non-Kathmandu studies using the chi-squared test for subgroup differences. Meta-regression examined three covariates individually and in a multivariable model: (i) publication year, (ii) log-transformed sample size, and (iii) geographic region. Pseudo-R^2^ was reported to indicate the proportion of between-study variance explained.

Publication bias was assessed visually with funnel plots and formally with Egger’s regression test. Influence diagnostics (Cook’s distance, hat values, DFBETAS) and leave-one-out (LOO) analysis were conducted to assess the robustness of key findings.

### Sensitivity analysis

A key source of heterogeneity is diagnostic method. The WHF echocardiographic criteria were published in 2012.[11] Studies conducted before 2012 (Shrestha 1991, Regmi 1997, KC Man Bahadur 2003) used auscultation or pre-WHF echocardiographic criteria, both of which are known to underestimate RHD prevalence compared with WHF-criterion echo screening. All three of these studies are from the Kathmandu subgroup. To assess whether the geographic disparity is partly attributable to this diagnostic confound, we note that the Kathmandu subgroup (pooled prevalence 1.1%) consists entirely of pre- or early-era studies, while the non-Kathmandu subgroup (pooled prevalence 7.2%) consists entirely of post-2012 WHF-criterion studies. The geographic comparison should therefore be interpreted with caution: observed differences partly reflect both true urban-rural disparities and the higher sensitivity of contemporary WHF-criterion echocardiography. A sensitivity analysis restricted to post-2012 studies using WHF criteria would require individual participant data or updated primary studies and is recommended as a priority for future research.

### Certainty of evidence

The GRADE framework, adapted for prevalence studies, was used to rate certainty across five domains: risk of bias, inconsistency, indirectness, imprecision, and publication bias. (12) Ratings were assigned as high, moderate, low, or very low.

### Software

All analyses were performed in R (version 4.3.2) using the meta and metafor packages. Statistical significance was defined as two-tailed p<0.05.

## Results

### Study selection

The search identified 70 records (67 from databases, 3 from other sources). After removing 20 duplicates, 47 unique records were screened. Of these, 38 were excluded at title and abstract stage. Nine full texts were retrieved; one hospital-based descriptive study (Joshi et al.) was excluded because the denominator was restricted to children with cardiac disease. (13) Two grey literature reports were assessed; one was excluded because the denominator comprised only patients with valvular heart disease (Basnet et al.). (14)Ten studies met all eligibility criteria and were included. The PRISMA flow diagram is presented as Figure 1.

**Figure 1.** PRISMA 2020 flow diagram of study selection.

### Study Characteristics

The ten included studies enrolled 209,815 participants in total, with individual sample sizes ranging from 2,795 to 107,340 and prevalence ranging from 0.9% to 13.2%. Four studies were conducted in the Kathmandu Valley (n = 53,732), five in non-Kathmandu regions (n = 48,743), and one was nationwide (n = 107,340). Table 1 summarises study characteristics.

**Table 1:**
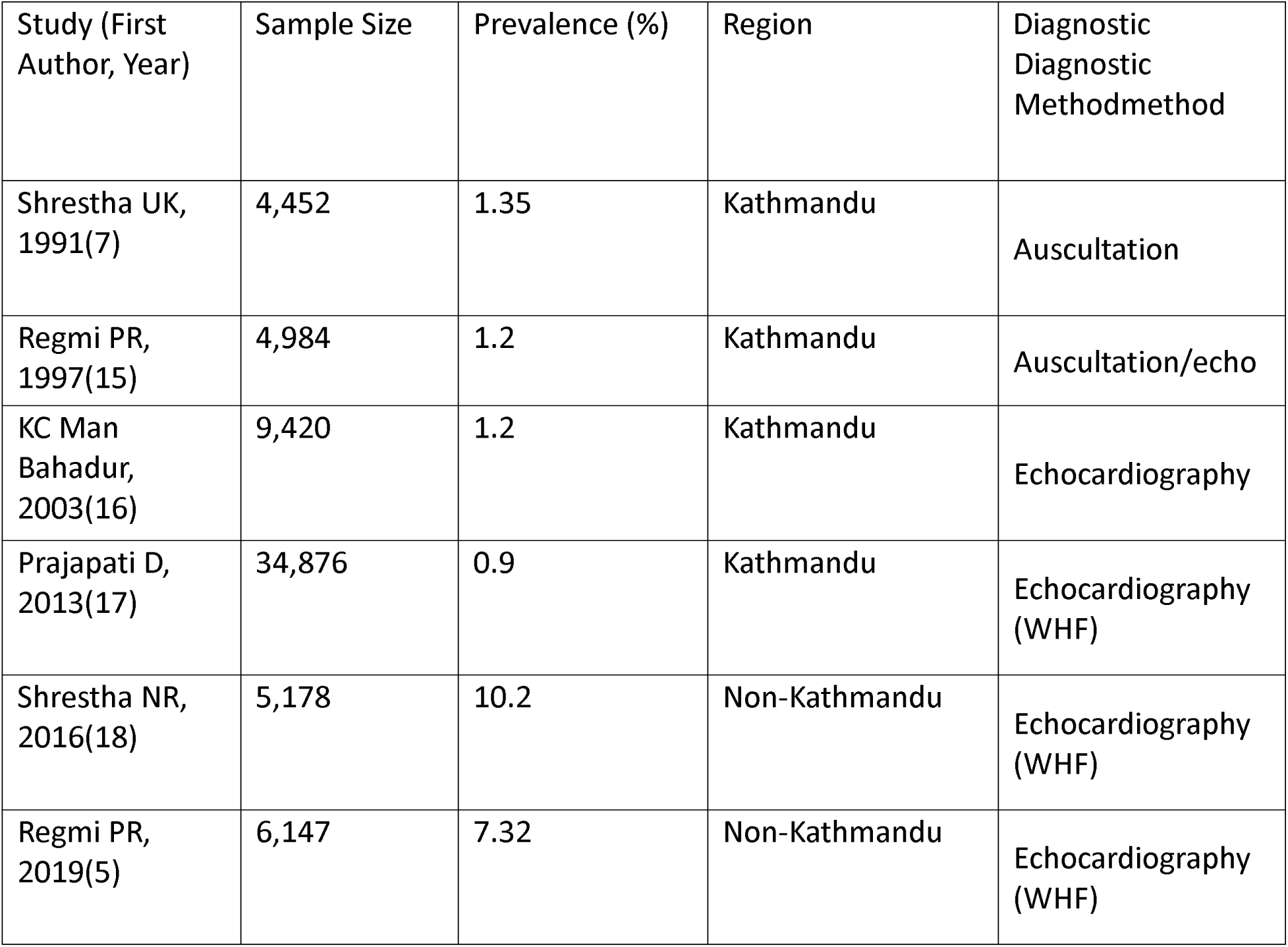
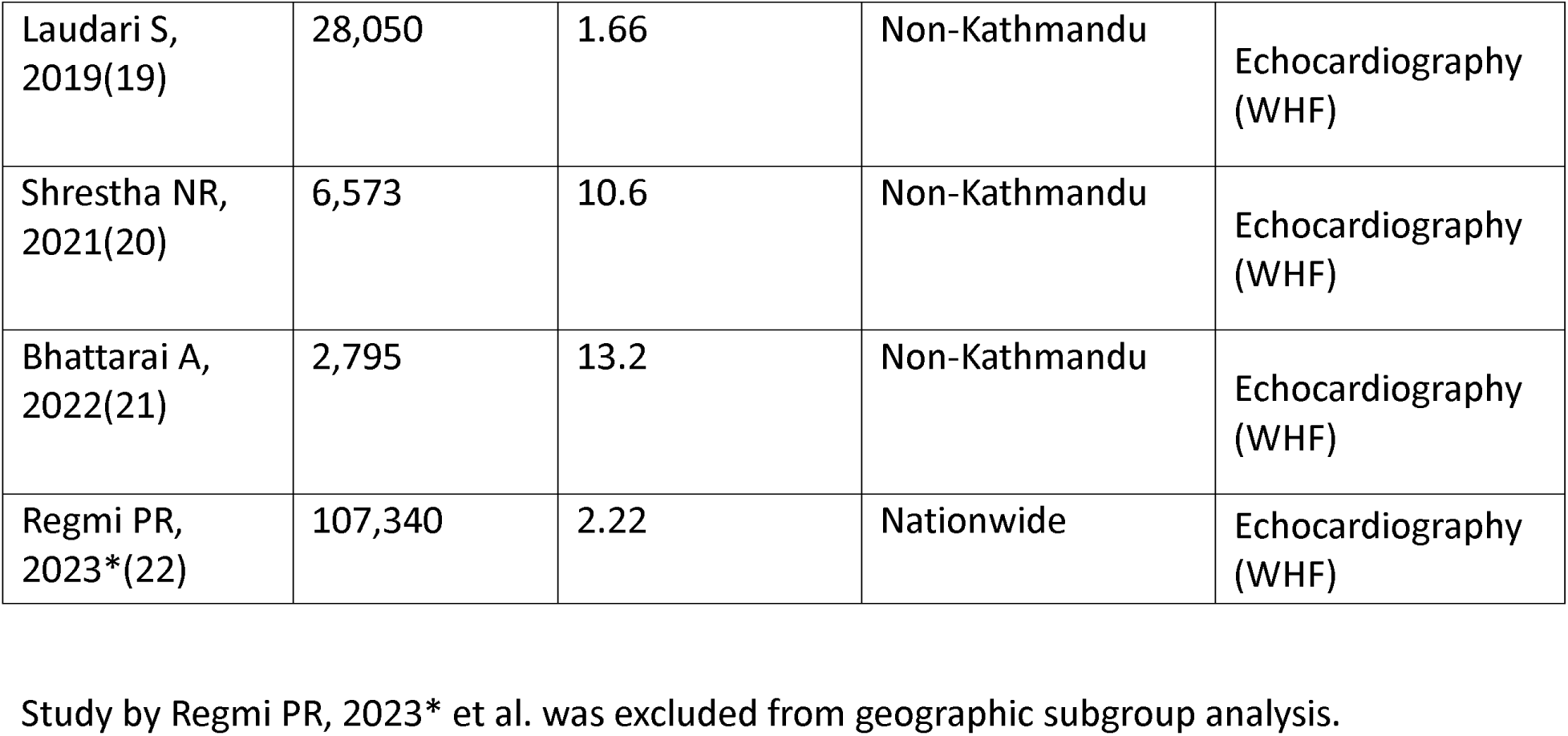
Characteristics of Included Studies.

### Risk of bias

Most studies employed appropriate sampling frames, recruitment strategies, and echocardiographic ascertainment. Sample size adequacy and response rate reporting were variable. Five studies were assessed as not meeting the JBI criterion for adequate sample size, and five had unclear response rates, contributing to the high heterogeneity observed. Table 2 presents the full JBI appraisal.

**Table 2.**
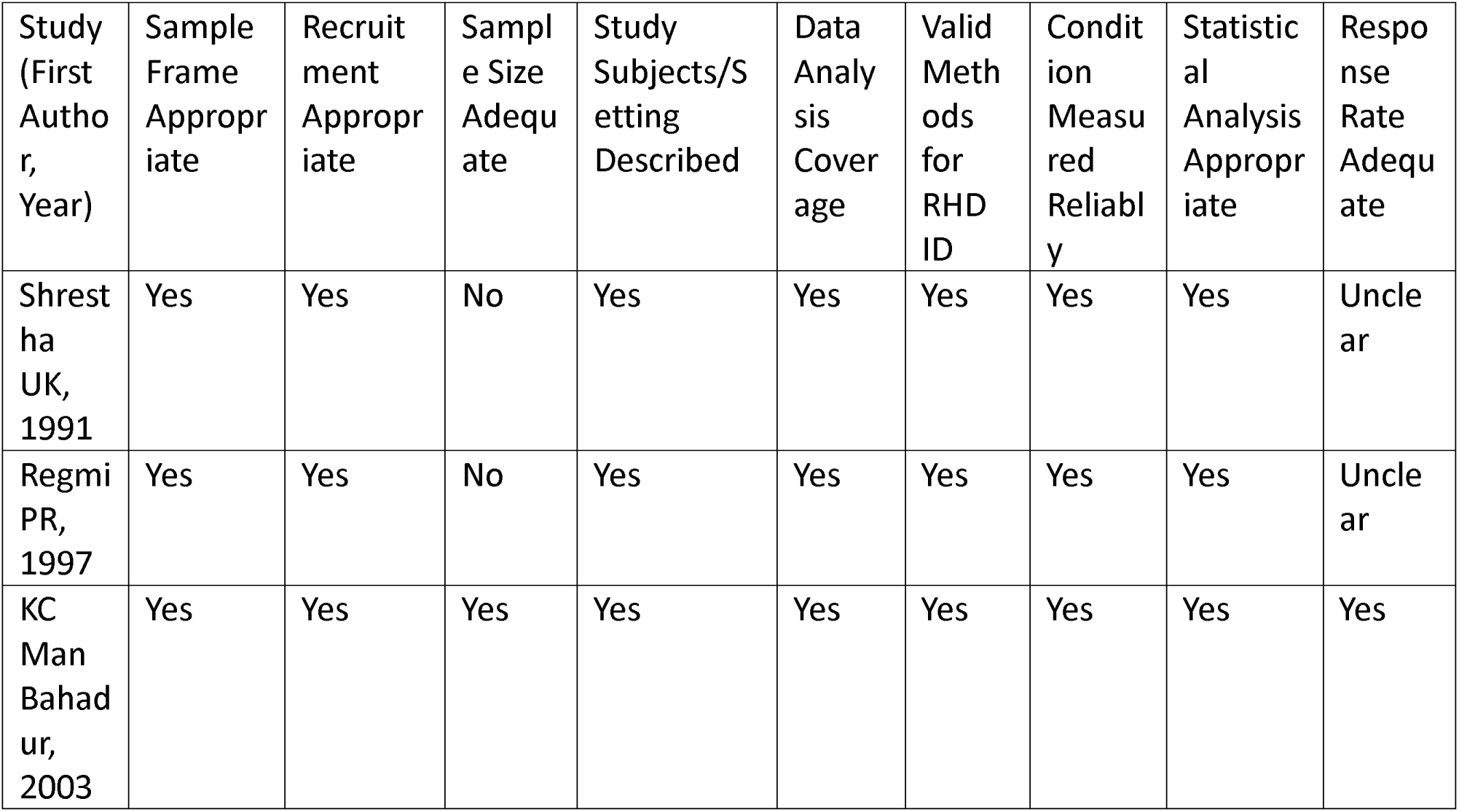
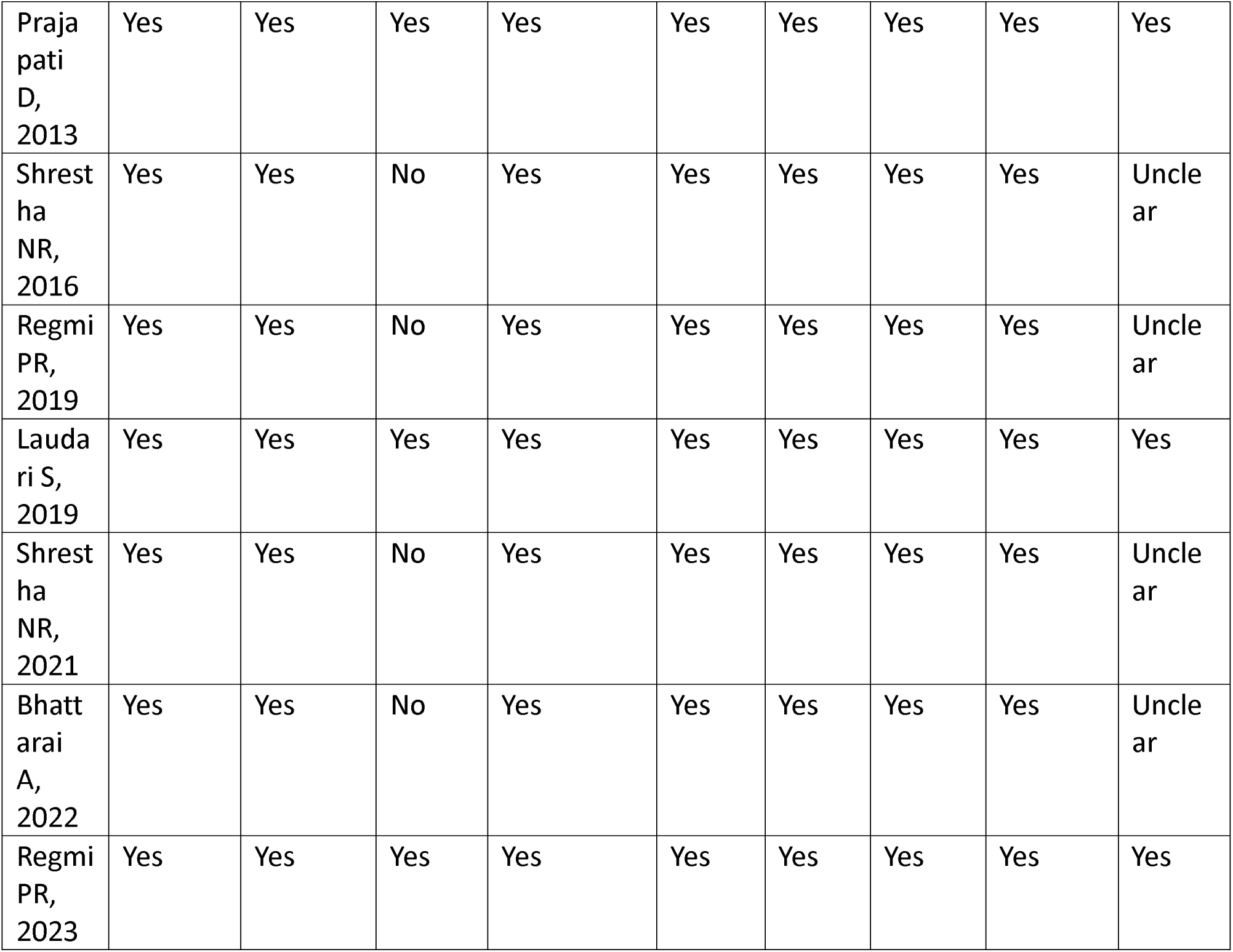
JBI critical appraisal of included studies.

### Prevalence

The overall pooled prevalence of RHD in Nepal was 3.1% (95% CI 1.6 to 5.8%). The prediction interval was 0.3% to 28.5%, indicating that the true prevalence in a new Nepalese setting could plausibly range from very low to very high. Heterogeneity was extreme (I^2^ = 99.8%; tau^2^ = 1.126; p<0.001). The forest plot is presented as Figure 2.

**Figure 2.** Forest plot of pooled RHD prevalence using a GLMM with logit transformation (random effects; back-transformed to %). The diamond represents the pooled estimate with 95% CI. I^2^ and tau^2^ quantify between-study heterogeneity. The prediction interval (0.3% to 28.5%) reflects the expected range of true prevalence in a new Nepalese setting.

### Geographic Disparities

Subgroup analysis revealed a six-fold difference in RHD prevalence between geographic groups. Pooled prevalence was 1.1% (95% CI 0.9 to 1.3%) in the Kathmandu Valley and 7.2% (95% CI 3.7 to 13.3%) in non-Kathmandu regions. The difference was statistically significant (odds ratio 6.52, 95% CI 4.83 to 8.81; p<0.001). Within-group heterogeneity was moderate for Kathmandu (I^2^ = 78.9%) and very high for non-Kathmandu (I^2^ = 99.7%). The subgroup forest plot is presented as Figure 3.

**Figure 3.** Subgroup forest plot by geographic category (Kathmandu Valley vs non-Kathmandu). The nationwide study (Regmi 2023) is excluded from subgroup analysis. p-value from chi-squared test for subgroup differences. Note that all Kathmandu studies used pre-WHF or auscultation-based diagnostic methods, while all non-Kathmandu studies used post-2012 WHF criteria; this diagnostic-era difference partially explains the observed disparity.

As noted in the methods, all Kathmandu studies were conducted before 2012 using auscultation or pre-WHF echocardiographic criteria, while all non-Kathmandu studies used post-2012 WHF criteria. This diagnostic-temporal confound means the geographic disparity estimate of 6.5-fold should be interpreted as an upper bound of the true urban-rural gap.

### Meta-regression

In univariable models, more recent publication year was associated with higher prevalence (beta = 0.072, 95% CI 0.016 to 0.128; p = 0.012), explaining approximately 40% of between-study heterogeneity. Larger sample size was associated with lower prevalence (beta = −0.87, 95% CI −1.19 to −0.56; p<0.001), explaining 32%. Non-Kathmandu region was associated with higher prevalence (beta = 1.82, 95% CI 1.35 to 2.29; p<0.001), explaining 49%.

In the multivariable model including all three covariates, publication year (beta = 0.063; p = 0.006) and log(sample size) (beta = −0.87; p<0.001) remained significant, while the regional coefficient attenuated to non-significance (beta = 0.49; p = 0.32). The model explained 93.5% of between-study heterogeneity (pseudo-R^2^ = 93.5%). This attenuation indicates that the apparent geographic contrast is partly explained by the fact that non-Kathmandu studies are both smaller and more recent. However, because the multivariable model includes only ten studies for three covariates, overfitting cannot be excluded and these coefficients should be treated with caution.

The temporal trend (rising prevalence with more recent year) does not reflect a true increase in RHD burden. Once sample size and region are controlled for, the temporal coefficient attenuates and the pattern is consistent with a combination of diagnostic upgrading (the shift from auscultation to WHF-criterion echocardiography) and small-study effects (smaller, high-prevalence, non-Kathmandu studies being published in more recent years). There is insufficient evidence to conclude that RHD burden has genuinely increased over the study period; equally, there is no evidence of meaningful decline. Table 3 presents the meta-regression results.

**Table 3.** Meta-regression results (logit scale)

| Covariate | Beta (95% CI) | p-value | Heterogeneity explained (R<sup>2</sup>) |
| --- | --- | --- | --- |
| Publication year (univariable) | 0.072 (0.016 to 0.128) | 0.012 | 40% |
| Log(sample size) (univariable) | -0.87 (-1.19 to -0.56) | <0.001 | 32% |
| Non-Kathmandu region (univariable) | 1.82 (1.35 to 2.29) | <0.001 | 49% |
| Multivariable: publication year | 0.063 (0.018 to 0.108) | 0.006 | Overall R<sup>2</sup> = 93.5% |
| Multivariable: log(sample size) | -0.87 (-1.19 to -0.55) | <0.001 |  |
| Multivariable: non-Kathmandu region | 0.49 (-0.49 to 1.47) | 0.32 |  |

CI = confidence interval; R<sup>2</sup> = pseudo-R-squared. Multivariable model includes publication year, log(sample size), and region simultaneously.

### Publication bias and influence analysis

Visual inspection of the funnel plot suggested modest asymmetry. Egger’s test was borderline (p = 0.096). Given the small number of included studies, this test is underpowered and results should be interpreted cautiously. Influence diagnostics identified Prajapati 2013, Regmi 2019, and Bhattarai 2022 as having disproportionate leverage. LOO analysis confirmed that pooled prevalence remained stable (range 2.7% to 3.4%) and that the directions of all meta-regression coefficients were preserved after sequential exclusion of each study. Figures 4, 5, and 6 present the bubble plot, funnel plot, and influence diagnostics respectively.

**Figure 4.** Bubble plot of RHD prevalence (%) versus publication year. Bubble size is proportional to sample size. The fitted line represents the multivariable meta-regression model (reference: non-Kathmandu = 0). The shaded band represents the 95% prediction interval.

**Figure 5.** Funnel plot of standard error versus residual effect size. Egger’s regression test p = 0.096. Given the small number of studies, the test is underpowered; results should be interpreted with caution.

**Figure 6.** Influence diagnostics (hat values, Cook’s distance, DFBETAS, tau^2^, QE, and weight) for the multivariable meta-regression model. Red points indicate influential studies. No single study reversed the direction of key associations.

### Certainty of Evidence

Using GRADE, overall prevalence was rated low certainty, downgraded for very high inconsistency (I^2^ = 99.8%; prediction interval 0.3% to 28.5%) and imprecision (wide 95% CI). Kathmandu prevalence was rated moderate, downgraded once for inconsistency across study years. Non-Kathmandu prevalence was rated low owing to very high heterogeneity and imprecision. The temporal trend was rated low to moderate; the direction was robust, but residual heterogeneity and small study number limited confidence. Table 4 presents the GRADE summary.

**Table 4.**
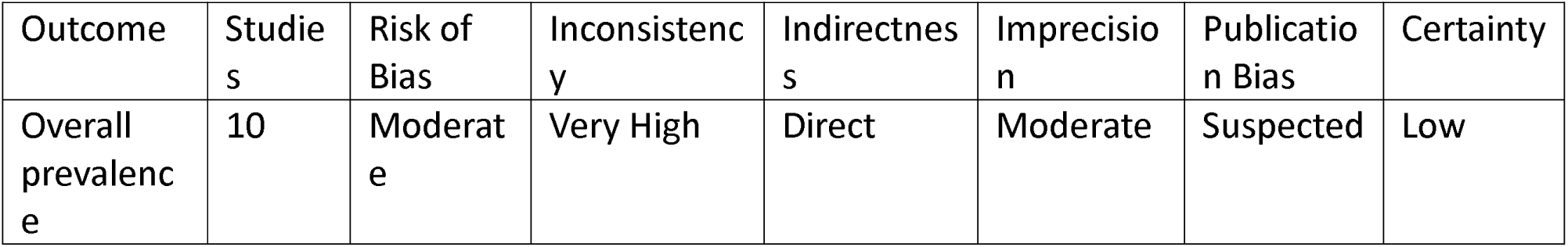
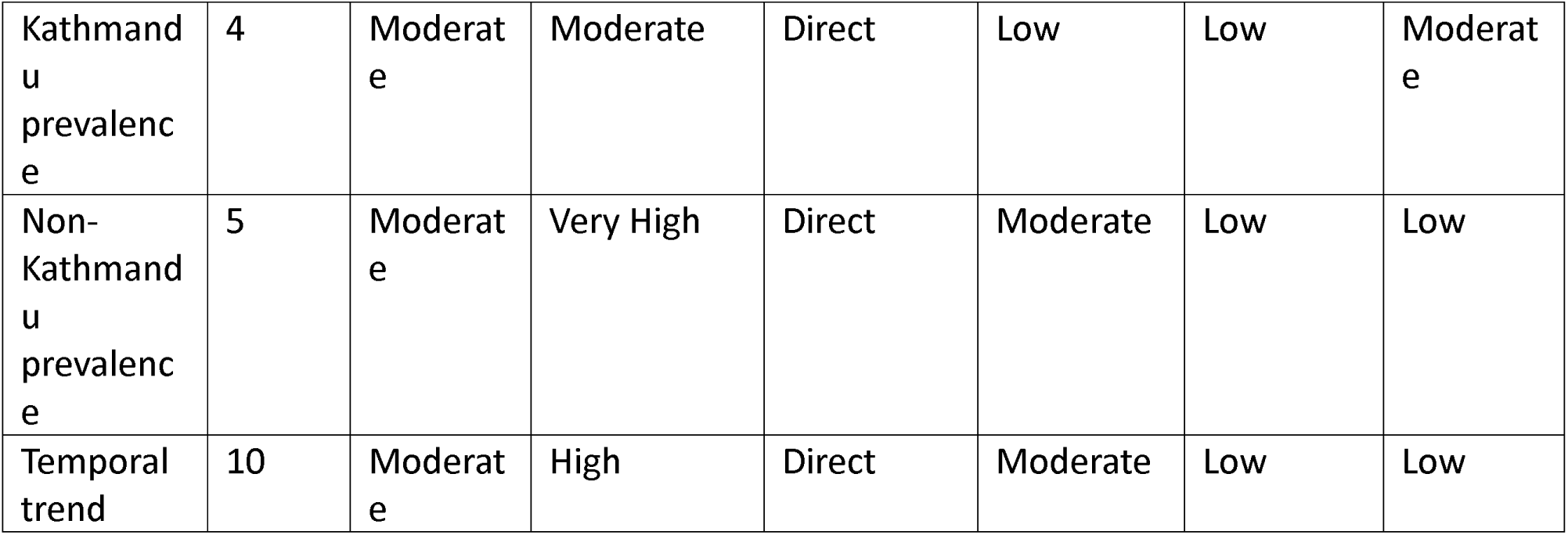
Certainty of Evidence (GRADE)

## DISCUSSION

### Summary of Findings

This meta-analysis, synthesising ten population-based studies and over 200,000 participants, provides the most comprehensive evidence to date on RHD burden in Nepal. The pooled prevalence was 3.1% (95% CI 1.6 to 5.8%), with a prediction interval of 0.3% to 28.5%, underscoring substantial heterogeneity. Prevalence in the non-Kathmandu group was 7.2%, approximately six times that of the Kathmandu Valley (1.1%). After accounting for sample size and diagnostic-era effects in multivariable meta-regression, the regional coefficient attenuated, and the temporal trend no longer held statistical significance. These findings collectively indicate that the RHD burden in Nepal is high, inequitably distributed, and has not demonstrably declined over three decades, though a definitive temporal conclusion is constrained by the diagnostic-era confound.

### Comparison with prior evidence

Our findings contrast with the global trajectory of RHD. A pooled analysis of echocardiographic studies found an overall global prevalence of 26.1 per 1000 using WHF criteria, with an inverse relationship with national income(23). In South Asia, a systematic review reported a 24.8% decline in prevalence from 1991 to 2021(8). Nepal presents a concerning deviation: the 2016 Eastern Nepal school-based study found RHD affecting 1 in 100 schoolchildren, with subclinical cases five times more common than clinically manifest disease(18). Despite global progress linked to improved living standards and benzathine penicillin access, Nepal’s community-level burden remains high (24).

The geographic disparity mirrors health system inequalities. The Kathmandu Valley has greater access to cardiologists, echocardiographic equipment, and referral pathways, alongside higher school attendance and health literacy. Outside Kathmandu, shortages of trained personnel, weak benzathine penicillin supply chains, inadequate school health programmes, household crowding, and seasonal migration sustain streptococcal transmission and compromise prophylaxis continuity. These structural determinants are consistent with global evidence linking socioeconomic deprivation to higher RHD burden(25). It is important to acknowledge, however, that the apparent six-fold gap is also partly an artifact of diagnostic-era differences: all Kathmandu studies used pre-WHF or auscultation-based methods, while non-Kathmandu studies used post-2012 WHF-criterion echocardiography. The true urban-rural gap may be smaller than our point estimate suggests.

### Strengths

It is important to acknowledge, however, that the apparent six-fold gap is also partly an artifact of diagnostic-era differences: all Kathmandu studies used pre-WHF or auscultation-based methods, while non-Kathmandu studies used post-2012 WHF-criterion echocardiography. The true urban-rural gap may be smaller than our point estimate suggests.

### Limitations

Several limitations must be acknowledged. Only ten studies were eligible, which constrains the precision of subgroup and meta-regression analyses. The multivariable meta-regression model includes three covariates for ten studies, raising the risk of overfitting; the 93.5% pseudo-R^2^ should not be interpreted as definitive. Geographic coverage was incomplete, with only three of Nepal’s seven provinces represented; prevalence may be even higher in unstudied remote regions. The diagnostic-era confound between Kathmandu (pre-WHF methods) and non-Kathmandu (WHF criteria) studies means the geographic comparison conflates location with diagnostic sensitivity. With only ten studies, Egger’s test for publication bias is underpowered, and the possibility of publication bias cannot be excluded. As an aggregate meta-analysis, individual-level determinants such as socioeconomic status, hygiene, and household crowding could not be examined.

### Policy Implications

The persistence of high RHD prevalence in Nepal has urgent implications across several levels. At the clinical level, systematic detection of latent RHD in school-aged children, coupled with timely initiation of benzathine penicillin prophylaxis, should be prioritised. At the health system level, integration of RHD control into Nepal’s decentralised federal structure is critical. Rural municipalities need reliable benzathine penicillin supply chains, training of frontline health workers in portable echocardiography, and community engagement to improve prophylaxis adherence. These efforts can be delivered within Nepal’s existing Basic Health Services Package and through ward-level health posts. Task-shifting to nurses and community health workers for echocardiographic screening has shown promise in comparable LMIC settings and merits formal evaluation in Nepal.[27] Linking RHD control to Nepal’s non-communicable disease strategy, which aligns with the WHO Nepal Country Cooperation Strategy, would create programmatic synergies and improve resource efficiency. At the upstream level, addressing the social determinants of RHD (poverty, overcrowding, poor sanitation) requires intersectoral action beyond the health sector.

### Future Research

Province-representative surveys using standardised WHF echocardiographic criteria in all seven provinces are needed to generate a reliable national prevalence estimate and identify geographic hot spots. A sensitivity analysis restricted to post-2012 WHF-criterion studies, using individual participant data, would allow a cleaner geographic comparison free of diagnostic-era confounding. Longitudinal studies tracking the natural history of screen-detected latent RHD under real-world prophylaxis conditions are required(26). Implementation research should evaluate the scalability and cost-effectiveness of portable echocardiography and task-shifting models in Nepal’s federal health system(27).

## Conclusion

RHD in Nepal is not only prevalent but profoundly inequitable. Prevalence outside the Kathmandu Valley is approximately six times higher than in urban populations, and there is no evidence of meaningful decline over three decades, though this conclusion is limited by diagnostic-era differences between studies. The combination of entrenched social determinants, weak health system capacity, and inconsistent prophylaxis threatens to perpetuate this burden. Addressing RHD in Nepal requires urgent investment in primary prevention, school-based echocardiographic screening, and equitable access to prophylaxis through the federal health system. This is both a medical and a social justice imperative.

## Data Availability

All data produced in the present study are available upon reasonable request to the authors

## Abbreviations

ARF: acute rheumatic fever
CI: confidence interval
GLMM: generalised linear mixed model
GRADE: Grading of Recommendations, Assessment, Development and Evaluation
I^2^: inconsistency index
JBI: Joanna Briggs Institute
LMICs: low- and middle-income countries
LOO: leave-one-out
OR: odds ratio
PRISMA: Preferred Reporting Items for Systematic Reviews and Meta-Analyses
PROSPERO: International Prospective Register of Systematic Reviews
RHD: rheumatic heart disease
tau^2^: between-study variance
WHF: World Heart Federation

## Declarations

### Ethics approval and consent to participate

Not applicable. This meta-analysis is based on the synthesis of previously published data and does not involve direct human or animal participation.

### Consent for publication

Not applicable. This manuscript does not contain any individual person’s data in any form.

### Availability of data and materials

All data analyzed in this meta-analysis are derived from publicly available studies. No new datasets were generated.

### Authors’ contributions

SP: conceptualisation; methodology; supervision; project administration; validation; writing (review and editing); guarantor. AT: investigation (screening, data extraction, risk-of-bias); data curation; validation; writing (review and editing). AB: investigation (database searches, screening); data curation; methodology; writing (review and editing). ATi: methodology (JBI appraisals); validation; data curation; visualisation; writing (review and editing). APa: data curation; investigation; visualisation; writing (review and editing). AN: formal analysis (GLMM, meta-regression); software; visualisation; methodology; writing (review and editing). DJS: methodology oversight; validation; supervision; writing (review and editing). KBB: writing (original draft); writing (review and editing). SD: resources; supervision; writing (review and editing). All authors reviewed and approved the final manuscript.

## Acknowledgements

Thankful for previous authors.

